# Frequency of pharmacogenomic variation and medication exposures among All of Us Participants

**DOI:** 10.1101/2024.06.12.24304664

**Authors:** Andrew Haddad, Aparna Radhakrishnan, Sean McGee, Joshua D. Smith, Jason H. Karnes, Eric Venner, Marsha M. Wheeler, Karynne Patterson, Kimberly Walker, Divya Kalra, Sara E. Kalla, Qiaoyan Wang, Richard A. Gibbs, Gail P. Jarvik, Janeth Sanchez, Anjene Musick, Andrea H. Ramirez, Joshua C. Denny, Philip E. Empey, All of Us Research Program Investigators

## Abstract

Pharmacogenomics promises improved outcomes through individualized prescribing. However, the lack of diversity in studies impedes clinical translation and equitable application of precision medicine. We evaluated the frequencies of PGx variants, predicted phenotypes, and medication exposures using whole genome sequencing and EHR data from nearly 100k diverse All of Us Research Program participants. We report 100% of participants carried at least one pharmacogenomics variant and nearly all (99.13%) had a predicted phenotype with prescribing recommendations. Clinical impact was high with over 20% having both an actionable phenotype and a prior exposure to an impacted medication with pharmacogenomic prescribing guidance. Importantly, we also report hundreds of alleles and predicted phenotypes that deviate from known frequencies and/or were previously unreported, including within admixed American and African ancestry groups.

## Introduction

The National Institutes of Health’s *All of Us* Research Program (*All of Us*) aims to accelerate precision medicine by recruiting one million diverse participants with a focus on those previously underrepresented in research(*1*). As of Winter 2024, >750,000 participants have been enrolled, >400,000 have contributed electronic health record (EHR) data, and nearly 250,000 have undergone whole genome sequencing (WGS). *All of Us* provides value to participants through continuous engagement and elective return of hereditary disease risk and pharmacogenomics (PGx) reports.

PGx can significantly impact drug efficacy and toxicity. Many commonly prescribed medications have genotype-guided dosing or drug selection recommendations in their Food and Drug Administration (FDA) approved labeling or in guidelines developed by the Clinical Pharmacogenetics Implementation Consortium (CPIC). There are >300 medications with PGx information in drug labeling and >25 guidelines which cover >100 gene-drug pairs with CPIC level A evidence with a guideline representing the highest level of evidence(*2*). Implementation of PGx in clinical practice has demonstrated reduced cardiovascular adverse events through use of genotype-guided antiplatelet therapy(*3, 4*). A multi-gene panel guided treatment has further demonstrated reduced adverse event (*5*).

Despite the strong evidence supporting the use of PGx data in clinical practice, broader implementation is limited in part by the relative paucity of data on the frequency of variants, alleles, and predicted phenotypes across diverse biogeographical groups. European populations are overrepresented in genetic research representing a vast majority of genome wide association studies to date(*6, 7*). Small sample sizes and the use of targeted genotyping assays or whole exome sequencing (WES) which are unable to interrogate every variant within PGx genes also limit characterization of rare variants and the discovery of novel haplotypes(*8, 9*). Further, while several studies have captured the incidence or prevalence of PGx medication prescribing among large health systems or using payer data, they have lacked the genetic data to estimate the true impact of PGx by linking medication exposure with clinically actionable variants for an associated gene in the same patients(*10–13*). Studies that have included genetic data have been limited in discovery due to integrated call sets generated from combining imputed array data with WES, which are unable to characterize all known PGx variants(*9, 14*). More recently, evaluation of over 200k UK Biobank participants demonstrated the utility of large cohorts in the discovery of novel haplotypes and estimation of known PGx allele and phenotype frequencies(*15*). However, it still lacked the diversity and the WGS data necessary to accurately estimate PGx allele and phenotype frequencies in all groups.

To characterize PGx genetic diversity, we report the frequency of PGx variants, alleles, and predicted phenotypes in 16 well-established PGx genes using WGS data from nearly 100,000 *All of Us* participants. Frequencies of variants were compared against existing reference databases and frequencies of rare variants and genotypes were identified. We then estimate the real-world impact of PGx by determining the proportions of participants who have potentially actionable findings; a predicted PGx phenotype and exposure to a medication with a prescribing recommendation from CPIC or the FDA. This study represents one of the largest and the most diverse PGx datasets with linked medication exposures events to date.

## Results

### Demographics

Table 1 shows the characteristics of the participants with available WGS data (n=98,590), EHR data (n=372,082), and those who had both WGS and EHR data (n=98,553). The average age of the participants was relatively similar across the three groups at 54.61, 54.02, and 54.61 respectively. The majority of the participants reported female sex at birth at 59.76%, 59.79%, and 59.79% respectively. Of the participants with available WGS data, the genetic ancestry groups with the highest frequency were the EUR, AFR, and the AMR genetic ancestry groups at 50.38% (49,668), 23.22% (22,897), and 16.12% (15,893) as presented in Figure 1A.

**Figure 1:**
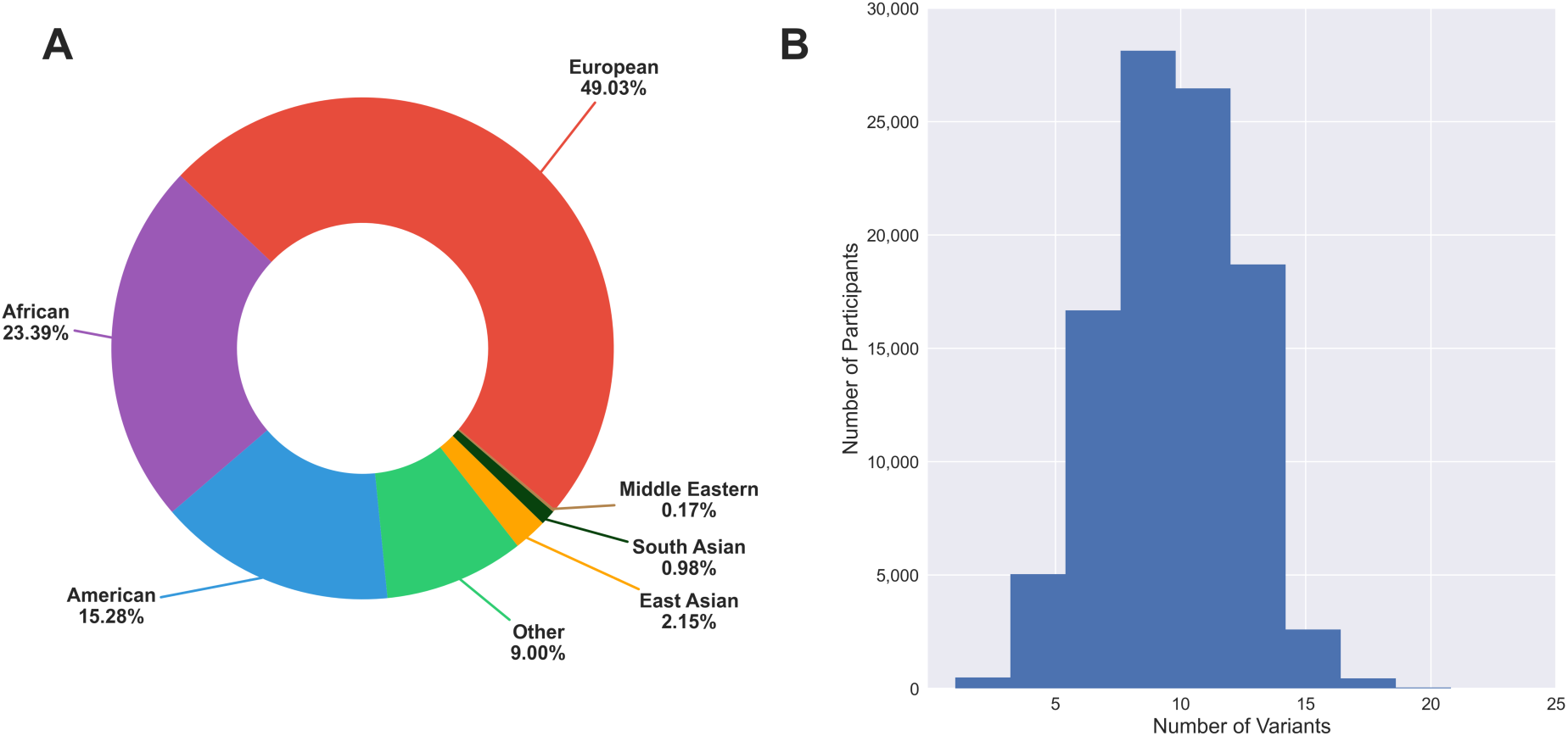
Genetic ancestry and variant distribution. Percentage of participants binned to each of the genetic ancestry super-populations (**A**). Distribution of the number of PGx variants observed in each participant (**B**).

**Table 1:** Participant demographics.

| **Characteristic** | **WGS** | **EHR** | **WGS + EHR** |
| --- | --- | --- | --- |
| **N** | 98,590 | 372,082 | 98,553 |
| **Age - mean (range)** | 54.61 (19-107) | 54.02 (19-122) | 54.61 (19-107) |
| **Sex at Birth - no. (%)** |  |  |  |
| - Female | 58,920 (59.76) | 222,463 (59.79) | 58,920 (59.79) |
| - Male | 38,133 (38.68) | 138,817 (37.31) | 38,133 (38.69) |
| - Intersex | 22 (0.02) | 79 (0.02) | 22 (0.02) |
| - Other/Missing | 1,515 (1.54) | 10,723 (2.88) | 1,478 (1.50) |
| **Imputed Sex - no. (%)** |  |  |  |
| - XX | 59,680 (60.53) | 59,652 (60.53) | 59,652 (60.53) |
| - XY | 38,167 (38.71) | 38,167 (38.71) | 38,167 (38.71) |
| - Other | 743 (0.75) | 743 (0.75) | 743 (0.75) |
| **Gender - no (%)** |  |  |  |
| - Female | 58,647 (59.49) | 220,800 (59.34) | 58,647 (59.51) |
| - Male | 37,970 (38.51) | 138,127 (37.12) | 37,970 (38.53) |
| - Non Binary | 160 (0.16) | 919 (0.25) | 160 (0.16) |
| - Transgender | 117 (0.12) | 464 (0.12) | 117 (0.12) |
| - Other/Missing | 1,760 (1.79) | 11,772 (3.16) | 1,659 (1.68) |
| **Genetic Ancestry - no (%)** |  |  |  |
| - European (EUR) | 49,668 (50.38) | 49,654 (50.38) | 49,654 (50.38) |
| - African/African American (AFR) | 22,897 (23.22) | 22,879 (23.21) | 22,879 (23.21) |
| - American Admixed/Latino (AMR) | 15,893 (16.12) | 15,889 (16.12) | 15,889 (16.12) |
| - East Asian (EAS) | 2,113 (2.14) | 2,113 (2.14) | 2,113 (2.14) |
| - South Asian (SAS) | 940 (0.95) | 940 (0.95) | 940 (0.95) |
| - Middle Eastern (MID) | 193 (0.20) | 193 (0.20) | 193 (0.20) |
| - Other (OTH) | 6,886 (6.98) | 6,885 (6.99) | 6,885 (6.99) |

### WGS variant analysis

WGS data for 955 variants was extracted for 98,590 participants. None of the variants were excluded due to low quality based on the *All of Us* Genomic Research Data Quality Report and no samples were dropped due to low call rate. Using a variant call rate cutoff of 0.98, five variants were dropped from any further analysis (*CYP2B6* c.329G>T, *CYP2B6* c.296G>A, *CYP2B6* c.785A>G, *SLCO1B1* c.1738C>T, and *NUDT15* c.156C>G). *CYP2B6* was dropped as a whole as the three dropped variants would have affected haplotype assignment for >50% of participants. Two variants (*ABCG2* c.421C>A and *NUDT15* c.7973C>T) from the AMR genetic ancestry group failed to meet HWE. Genotype concordance between WGS and genotyping array for these two variants across all samples (n=95,596 overlapping) and within the AMR group (n=14,601 overlapping) were >99%. Therefore, these variants were included in the remaining analysis as the deviation from HWE is unlikely to be due to genotyping error. Overall, every participant carried at least one PGx variant with a median of nine (Figure 1B). Variant frequencies overall and by genetic ancestry are presented in table S1.

### PGx Haplotyping

Stargazer and orthogonal analysis using PharmCAT produced very similar results disagreeing on only 0.12% (1,819/1,478,850) of all genotype calls. These differences were in *CYP2C19* (n=949), *SLCO1B1* (n=697), *VKORC1* (n=96), *G6PD* (n=59), *UGT1A1* (n=7), *ABCG2* (n=5), *CYP2C9* (n=3), *DPYD* (n=2) and *CYP3A5* (n=1). Importantly, most (64.65%; 1,176/1,819) of the differing genotype calls resulted in the same predicted phenotype. Therefore, the overall difference in the predicted phenotypes was only 0.04% (643/1,478,850). No notable differences were observed due to genetic ancestry. Stargazer and PharmCAT allele frequencies overall and split by genetic ancestry are reported in table S4. Similar analyses of phenotype frequencies are reported in table S5.

Using Stargazer, 99.48% (1,471,158/1,478,850) of gene/gene region calls were successfully assigned a genotype. Similarly, excluded variants resulted in *SLCO1B1* *45 and *46 calls being called as *1 and *15 respectively and *NUDT15* *12 was called as *1. The primary reason for an inability to assign a single genotype was due to a participant carrying >2 alleles which represented 81.23% (6,248/7,692) of the unsuccessful calls. Other reasons were an inability to distinguish between more than one possible genotype (13.55%; 1,042/7,692) and an inability to assign a genotype call based on the composite of variants identified in a given sample (did not meet any known core allele definitions) (5.23%; 402/7,692). A small (1.53%; 22,573/1,478,850) proportion of genotype calls returned >2 alleles (>1 allele for *G6PD* XY samples) for a gene suggesting incomplete haplotype definitions for these genes. Of the 98,590 participants, 16.50% (16,264/98,590) and 5.94% (5,854/98,590) had >2 alleles for *DPYD* and *CYP4F2* respectively. All genotype counts with >2 alleles are presented in table S6. Once *CYP4F2* is excluded due to a recent PharmVar update which added a novel *4 allele that is the combination of *2 and *3, 24.03% (5,501/22,897) of participants in the AFR group had at least one gene with >2 alleles, representing 40% more participants than the next highest group of MID at 17.10% (33/193).

*SLCO1B1* indeterminate phenotypes were found to be unusually high in the AFR group at 22.83% (5,227/22,897; Figure 2) primarily the result of relatively common *27, *41, and *43 alleles (4.78%, 3.20%, and 2.59% respectively) whose function has not yet been characterized versus allele frequencies that are ≤0.006 (table S4) in all other groups (excluding OTH). Similar results were observed for *TPMT* indeterminate phenotypes in the AFR group at 9.19% (2,105/22,897) due to undefined function for *8 and *24 alleles (2.54% and 2.35% respectively).

**Figure 2:**
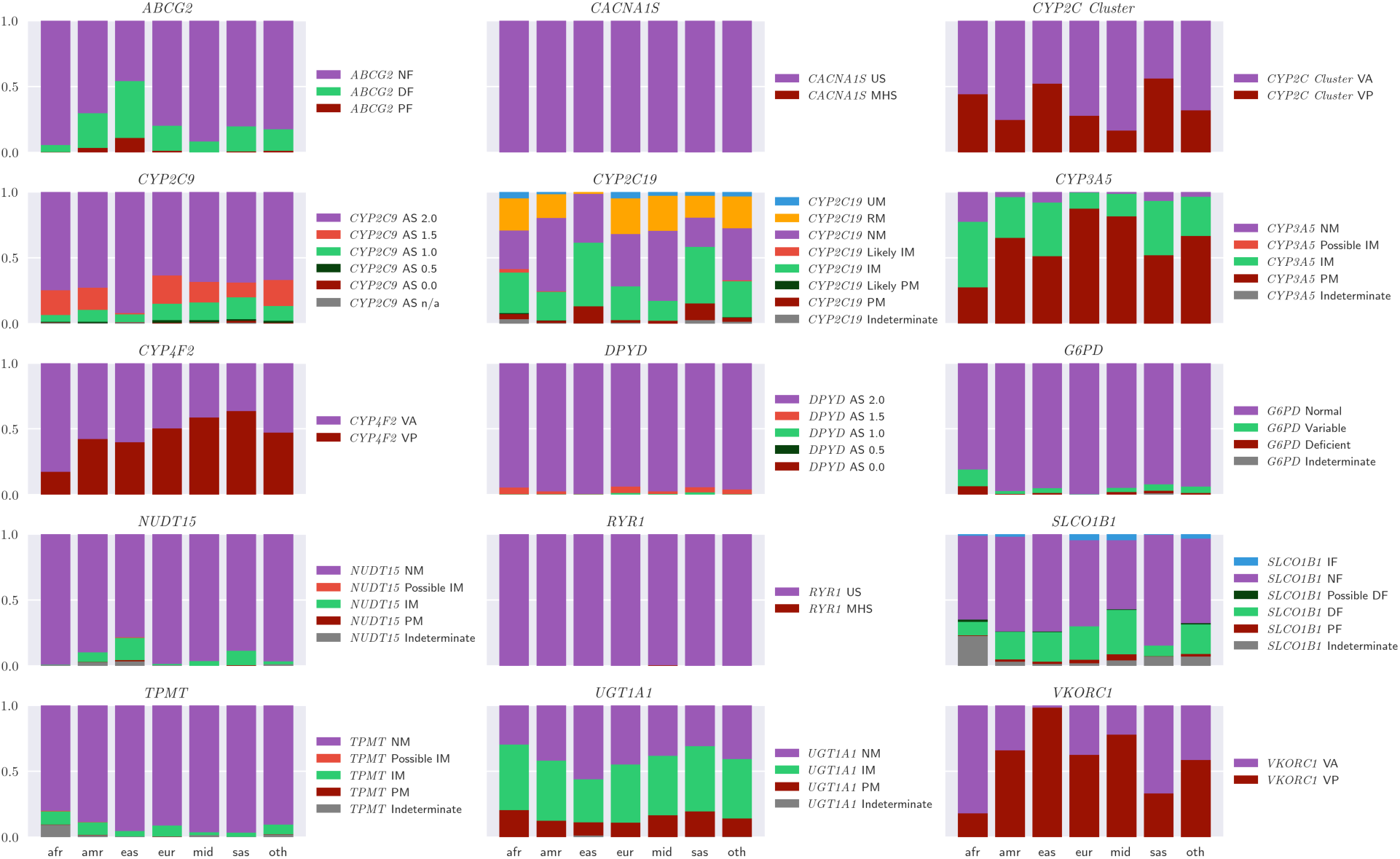
Phenotype frequency by genetic ancestry. NF: Normal Function, DF: Decreased Function, PF: Poor Function, AS: Activity Score, VA: Variant Absent, VP: Variant Present, NM: Normal Metabolizer, IM: Intermediate Metabolizer, PM: Poor Metabolizer, US: Uncertain Susceptibility, MHS: Malignant Hyperthermia Susceptibility, UM: Ultrarapid Metabolizer, RM: Rapid Metabolizer

Most (98.18%; 96,791/98,590) participants carried an actionable phenotype for at least one gene and 87.98% (86,744/98,590) carried an actionable phenotype for at least 2 genes based on CPIC guidelines. This expands to 99.13% (97,734/98,590) for at least 1 gene and 93.29% (91,971/98,590) for at least 2 genes when expanding to include FDA recommendations. This effect is caused by the *UGT1A1* intermediate metabolizer phenotype which is considered actionable for irinotecan by the FDA and does not yet have a guideline in CPIC. Actionable phenotypes were most commonly observed in UGT1A1 (59.25%; 58,413/98,590), CYP2C19 (59.10%; 58,264/98,590), and VKORC1 (52.93%; 52,186/98,590). The overall frequency of actionable phenotypes was high and similar across ancestry groups, ranging from 97.55% in the AMR group to 99.91% in the EAS group. However, there are notable differences between genetic ancestry groups in the frequency of actionable phenotypes within individual genes such as with *ABCG2* which was substantially more common in the EAS group at 10.84% versus the AFR group at 0.07% as shown in Figure 3. Similarly, most participants in the AFR group (72.44%) carried an actionable phenotype for *CYP3A5* compared to the EUR group (12.70%).

**Figure 3:**
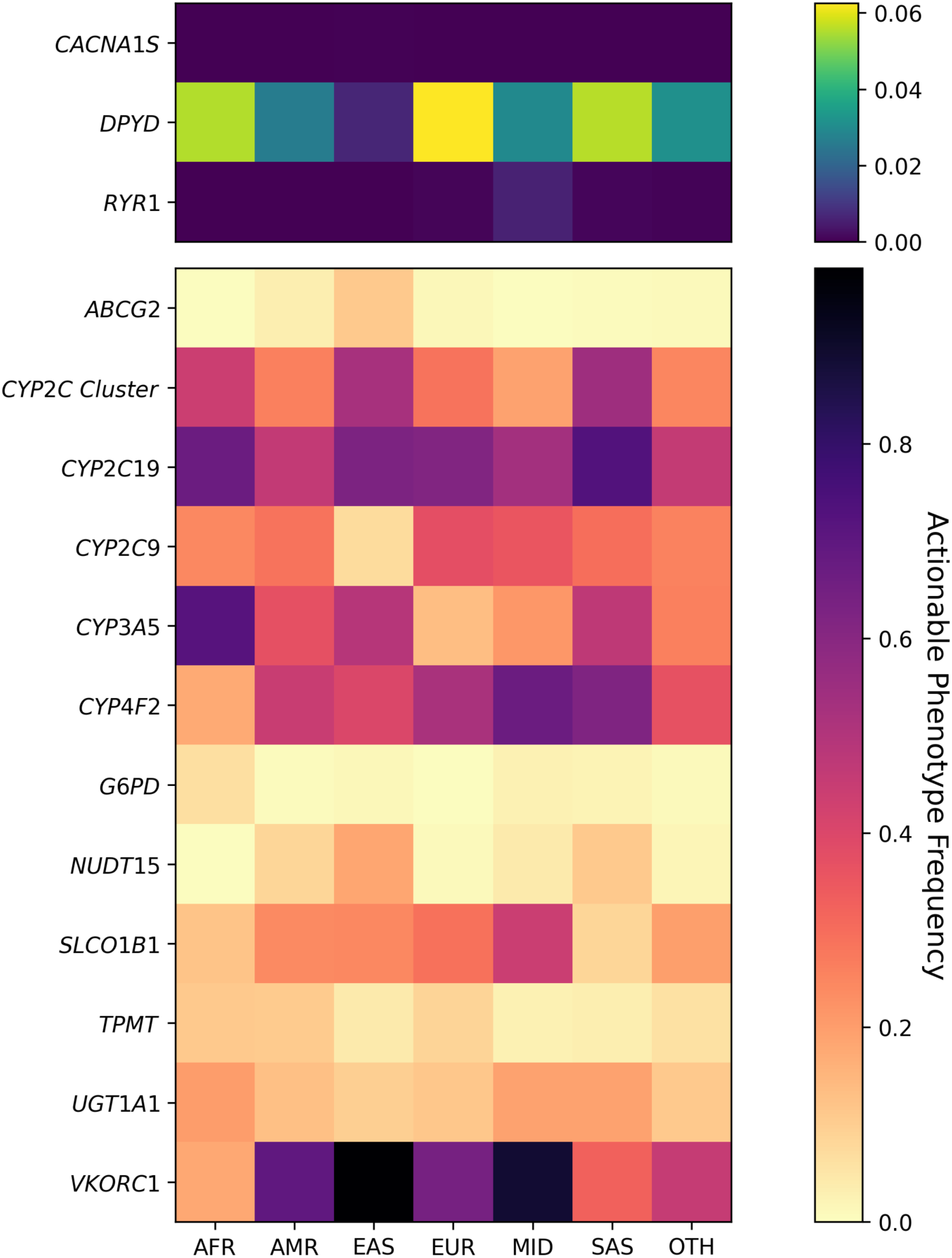
Frequency of actionable phenotypes. Predicted phenotypes were used to identify the frequency of actionable phenotypes by gene and genetic ancestry.

Although phased data was not available at this time, *UGT1A1* *37 was never observed alone in a sample; it was always in combination with the *80 allele. Further, *27 was observed only in the presence of both *28 and *80, likely representing a single haplotype. This could explain 31/34 *UGT1A1* genotype calls with >2 alleles in table S6. *CYP2C19* *2 and *30 were also commonly observed together and representing 28/163 instances of participants carrying >2 alleles for *CYP2C19*. Similarly, *TPMT* *8 and *33 represent all (n=11) instances of multiple alleles observed for this gene. Additional novel combinations that help to explain samples with >2 alleles include *SLCO1B1* *4 +*27, *26+*37 and *CYP2C9* *5+*36, *9+*36, *9+*11.

### Frequency comparisons

Frequencies were compared within genetic ancestry groupings to external, gold standard repositories (gnomAD and CPIC) whenever variants, non-reference alleles, and phenotypes have a known frequency >0. The OTH group was excluded as it is a heterogeneous population without a mapped comparator. Allele and phenotype frequencies were not compared for *CACNA1S* and *RYR1* due to the rarity of finding variants within these genes. Variant frequencies were similar to gnomAD with only 5.85% (58/992) of the variant-genetic ancestry comparisons being below the p-value cutoff and that most (35/58) were within the AMR group (Fig 4A; table S7). However, absolute differences in the frequencies were minimal, typically (55/58) less than 2%. Of the available biogeographical allele frequencies within CPIC, 25.75% (111/431) did not match *All of Us* genetic ancestry allele frequencies (Fig 4B; table S7). The AFR (n=41) group had the most alleles where the frequencies were significantly different from CPIC, whereas the AMR, EAS, MID, and SAS groups had much fewer allele frequencies which did not match CPIC at 10, 10, 2, and 10 respectively. Further, 35.78% (73/204) of the phenotype comparisons failed to match the respective biogeographical group frequencies within CPIC (Fig 4C; table S7). 41/73 differing frequencies were found to be significantly higher in *All of Us* than previously reported. The AFR genetic ancestry group was significantly different than reported CPIC frequencies for every phenotype for *SLCO1B1*, *TPMT* and *UGT1A1*. For example, the *SLCO1B1* decreased function phenotype was found to be at 10.38% (2,351/22,897) in *All of Us* for the AFR group while it was reported as 1.98% within CPIC in the African/African American biogeographical group.

**Figure 4:**
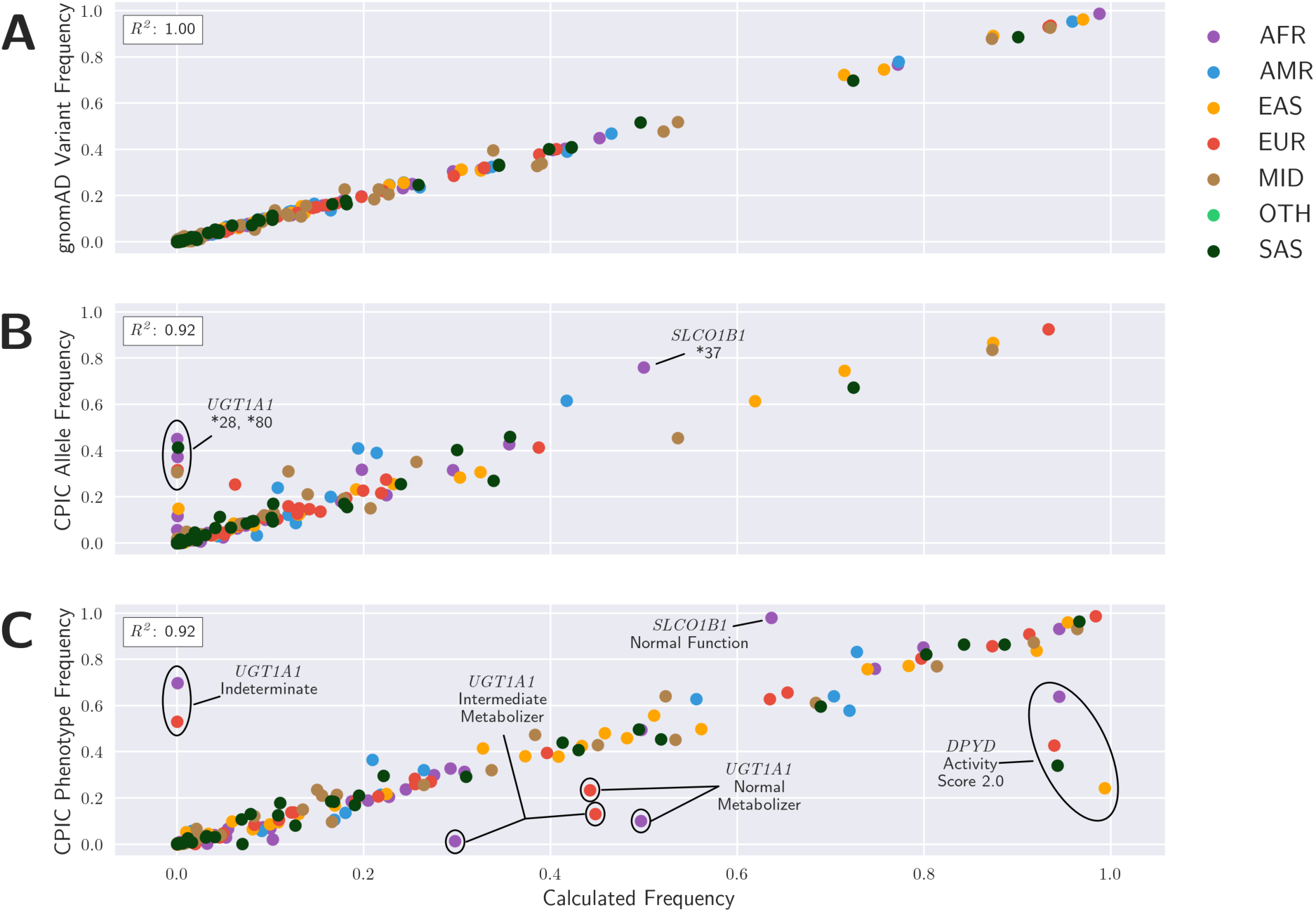
Comparison of calculated frequency and expected frequencies. Calculated variant frequencies compared to gnomAD (**A**). Allele (**B**) and phenotype (**C**) frequencies compared to CPIC.

### Identification of unknown frequencies

Overall, 491 allele and 129 phenotype frequencies by genetic ancestry groups (excluding OTH) were reported for the first time (these alleles and phenotypes were previously unknown or reported as “0” for a frequency within CPIC) per table S8 and Figure 5. The AMR and AFR groups had the highest number of previously unknown allele frequencies at 33.60% (165/491) and 24.25% (124/491) respectively. Fewer novel frequency estimates were made in the MID and SAS populations at 8.15% (40/491) and 5.70% (28/491) respectively; likely due to more limited sample size within this initial *All of Us* data release.

**Figure 5:**
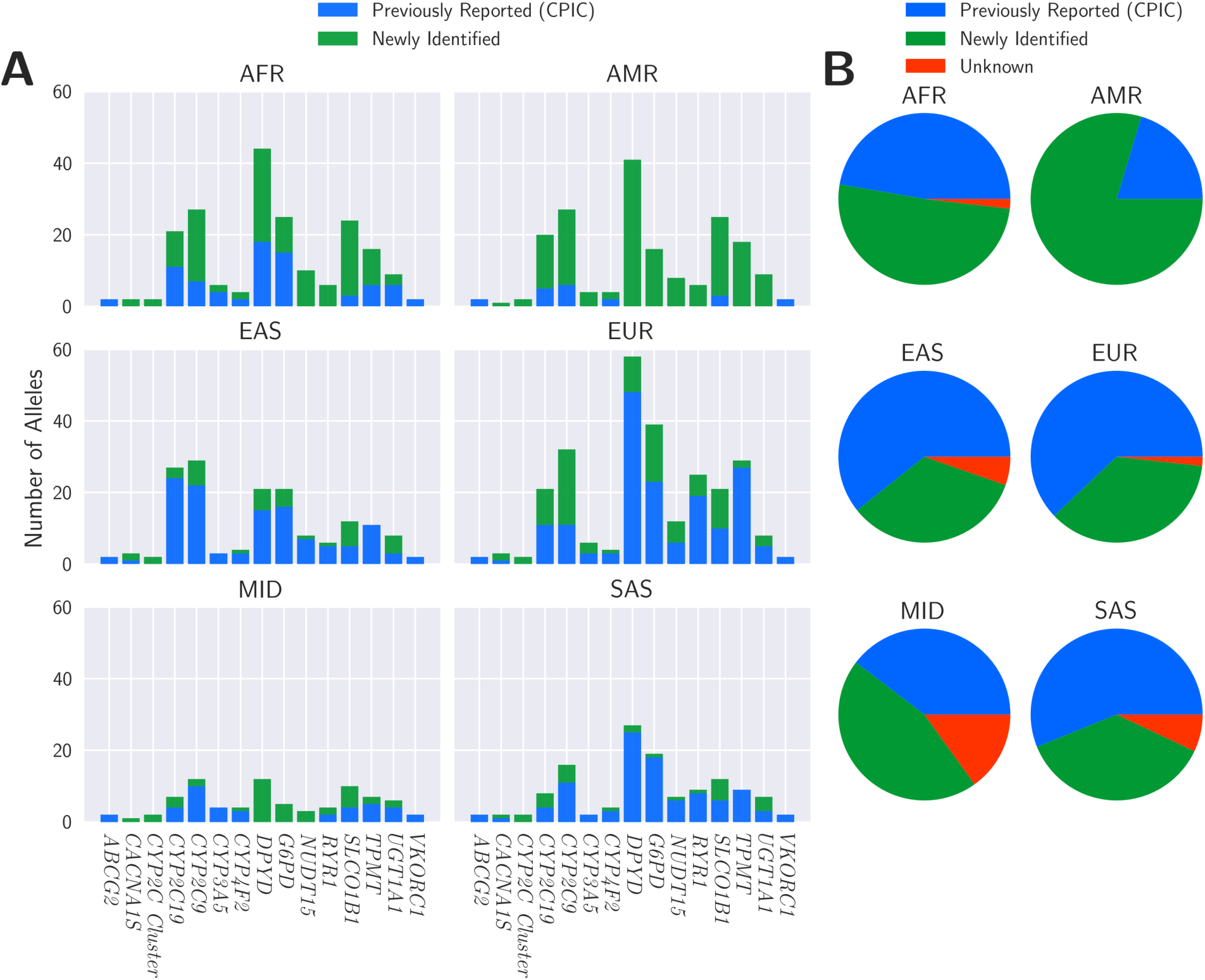
Newly computed allele and phenotype frequencies. Allele (**A**) and phenotype (Fig **B**) frequencies that were previously unknown or reported as 0 (Green). Frequencies previously known (Blue) and phenotype frequencies that remain unknown (Red). Allele frequencies that are unknown are not provided as the true number of possible alleles in each genetic ancestry population is unknown.

### PGx medication exposures among All of Us participants

EHR data was extracted for 372,082 participants with 68,576,017 medication exposure events. Participants had an average of 184 medication exposure events. Many (45.91%;170,829/372,082) participants had exposure to at least 1 medication with CPIC A guidance with a range of 0-24. When including additional medications that had FDA PGx guidance, 47.27% (175,895/372,082) of participants had an exposure to at least one PGx medication with a range of 0-29. The most common PGx medication exposures include ondansetron, ibuprofen, and omeprazole at 27.80% (103,434/372,082), 20.07% (74,683/372,082), and 14.15% (52,643/372,082) as shown in Figure 6. table S9 represents the overall prevalence of all PGx medications.

**Figure 6:**
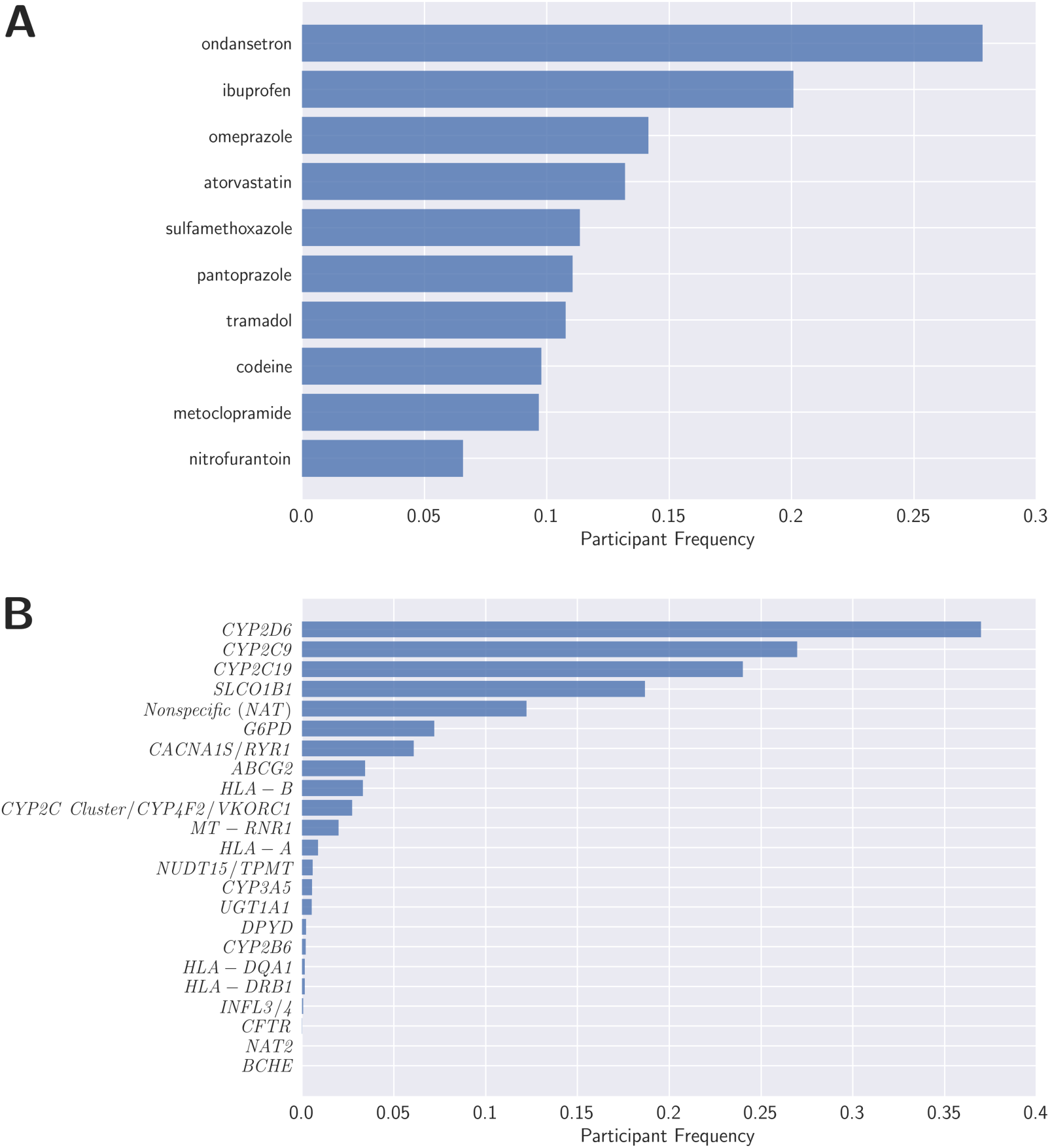
Prevalence of PGx medication exposures. Medication exposure prevalence overall for all PGx medications and for each of the top 10 most common medications (**A**). Prevalence of genes associated with medication exposures (**B**).

### Medication exposure with actionable phenotype

Of the participants who had both WGS and EHR data, 20.74% (20,442/98,553) carried an actionable phenotype for a medication that was part of a CPIC level A gene-drug pair which they have previously been exposed to. This rises only slightly to 20.80% (20,500/98,553) when expanding to include FDA PGx medications. When considering only CPIC medications, the most common PGx medication exposures in participants with an actionable phenotype for at least one associated gene were omeprazole, pantoprazole, and atorvastatin at 10.11% (9,960/98,553), 7.58% (7,473/98,553), and 3.77% (3,712/98,553) respectively as shown in Figure 7B. The genes with the most common actionable phenotype with an associated PGx medication exposure were *CYP2C19*, *SLCO1B1,* and *CYP2C9* at 15.09% (14,878/98,553), 4.68% (4,617/98,553), and 2.78% (2,736/98,553) respectively as shown in Figure 7A.

**Figure 7:**
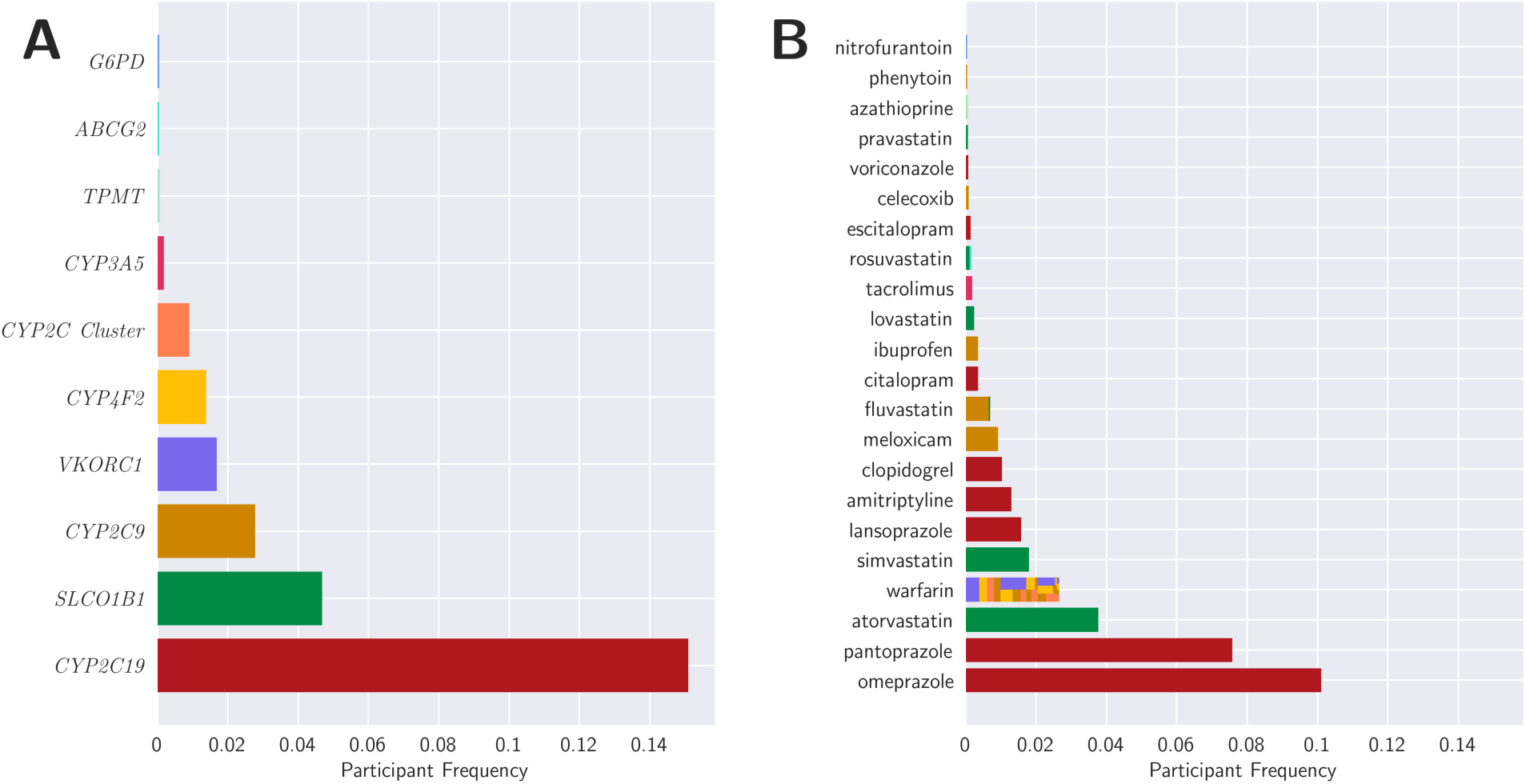
PGx medication exposures in participants with actionable phenotypes. Frequency of genes with actionable phenotypes associated with medication exposures (**A**). Frequency of a medication exposure while also carrying an actionable phenotype for an associated gene (**B**). Colors represent the associated gene for which the participant carries an actionable phenotype.

## Discussion

*All of Us* provides a unique opportunity to evaluate the frequency of PGx variants, actionable phenotypes, and associated medication exposures in one of the largest and most diverse cohorts with available WGS and EHR data. In the current analysis, we report all known variant, allele, and phenotype frequencies from 98,590 diverse participants across 15 pharmacogenes (excludes *CYP2B6*). We report 100% of participants carried at least one PGx variant and nearly all (99.13%) had a predicted phenotype for a gene which has a recommendation for a change in prescribing. Many allele (26%) and phenotype (36%) frequencies were found to deviate significantly from gold standard databases. A significant number of allele (491) and phenotype (129) frequencies for specific genetic ancestries have been estimated for the first time. Similarly, many novel haplotypes from known allele combinations are reported, demonstrating the value of a large diverse *All of Us* cohort with WGS data. Importantly, over 20% of participants had an actionable phenotype and previous exposure to a medication with CPIC or FDA PGx prescribing guidance for that phenotype, emphasizing the broad, real-world value of PGx.

The comparison of calculated frequencies to known gold standard databases represents opportunities to evaluate existing frequencies and determine where gaps exist in current knowledge. Variant frequency comparisons to gnomAD matched well with most of the differences representing minimal absolute differences. Comparisons to CPIC, however revealed larger differences, particularly in *SLCO1B1* and *UGT1A1* as well as the AFR and AMR groups where most allele and phenotype frequencies did not match CPIC. It is important to note that CPIC computes diplotype frequencies using data from studies with available allele frequencies based on HWE principle and then computes phenotype frequencies from the computed diplotypes. This process has the potential to expand the margin of error within the data. These previously established allele and phenotype frequencies are also negatively impacted due to use of targeted PGx assays or exome data that may not interrogate all variants, small sample sizes that are not capable of accurately capturing the frequency of rare alleles, and limited diversity in prior studies(*16*). Collectively, this has resulted in many allele and phenotype frequencies that are either unknown or reported as 0 which limits the number of comparisons that are possible. This can be clearly observed in table S3 where only 14 allele and 16 phenotype frequency comparisons (out of 185 and 54 respectively) be made to CPIC for the AMR group. The EAS, MID, and SAS groups appeared to match CPIC frequencies better, however this is likely attributed to the more limited numbers in this early release of *All of Us* data that better match the studies used to calculate the known frequencies within CPIC. The *All of Us* cohort represents one of the largest cohorts of participants from African ancestry at nearly 23k participants at the time this study was performed. The size of the cohort allowed for accurate estimation of allele frequencies and represented the group with the highest number of allele frequencies (n=41) that deviated from CPIC. The most notable frequency discovery is that every calculated phenotype frequency for the AFR group for *SLCO1B1*, *TPMT*, and *UGT1A1* was significantly different than in CPIC. This further highlights the underrepresentation of participants from African ancestry in genomic studies as a whole(*6*).

*All of Us* has particular value in understanding the frequency of rare variants because of the size of the cohort, comprehensiveness in genotyping approach, and the genetic diversity. Large biobank analyses demonstrate value to the field through improved understanding of haplotype frequencies which guides allele selection on clinical tests, aids in quality control for existing tests and allows clinical teams to estimate the impact of a panel design. In a recent UK Biobank analysis of over 200k participants, the frequency of 430 allele frequencies were computed for the first time compared to 491 values within this *All of Us* analysis(*15*). First, in UK Biobank many of the novel frequencies were computed in *CYP2B6* and *CYP2D6*. In this *All of Us* analysis, we did not report *CYP2B6* haplotypes because of stringent quality control. *CYP2D6* was not characterized because the initial release of genetic data did not include CRAM files that are needed to accurately characterize structural variants and haplotypes. Although *CYP2D6* allele frequencies were computed in UK Biobank, these data would not represent accurate haplotype frequencies without structural variant characterization. Exclusion of CYP2C Cluster allele frequencies to match values not included within the UK Biobank analysis gives a true gene to gene comparison of 285 vs 479. Despite *All of Us* having less than half the number of participants of the UK Biobank analysis at the time of these analyses, the difference clearly demonstrates the lack of knowledge on diverse cohorts and the power of having both expanded diversity with WGS data within *All of Us*. The computed frequencies provide further knowledge to improve equitable application of precision medicine through improved guidance of PGx clinical testing.

Many genotype calls suggested a sample was carrying >2 alleles. These situations represent important opportunities to update known haplotype definitions. *DPYD* genotypes could have more than two alleles due to the length of the gene that results in haplotype blocks instead of distinct haplotypes(*17*). *CYP4F2* has previously been shown in phased data that samples can carry the *2 and *3 variants on a single strand(*9, 18*) and has recently been updated by PharmVar with a new *4 allele which is defined as the combination of the *2 and *3 alleles. *UGT1A1* haplotypes have not yet been characterized by PharmVar despite the presence of a CPIC guideline and therefore leaves the potential for novel combinations of variants/alleles as was observed in the *80+*28+*27 haplotype(*19*). The *UGT1A1* *80+*28+*27 and *CYP2C19* *30+*2 haplotypes observed in *All of Us* participants were also recently identified among UK Biobank participants(*15*). *TPMT* *8+*33 has previously been described by GeT-RM in a trio as a novel *46 allele, however this allele definition has not yet been incorporated into CPIC but is recognized by the *TPMT* nomenclature committee(*20, 21*). The remaining combinations represent novel haplotypes and present an opportunity to update allele definitions within CPIC and PharmVar for these genes to better represent the haplotypes that are observed in diverse populations(*22*). Samples where a genotype call could not be assigned due to the combination of variants that were observed can also be identified from the dataset and represent additional opportunities for improved haplotype characterization.

Haplotype calling using Stargazer and PharmCAT demonstrated very high genotype and predicted phenotype concordance. Both tools attempt to resolve the genotype call for a gene by matching identified variants with known allele definitions. The small differences observed may be attributable to the assumptions that each tool makes about input data contribute to minor calling differences. The first difference is that PharmCAT considers suballeles as possible allele assignments whereas Stargazer does not. PharmCAT considers no calls for variants as a possible variant whereas Stargazer assumes the variant is homozygous reference. In genes where there is only a single variant defined by CPIC and the variant is not called for the sample, PharmCAT is unable to assign a genotype whereas Stargazer assigns the reference allele. With unphased data such as in the current analysis, both tools assume all variants are on the same chromosome strand (cis conformation). However, if all variants cannot be phased to the same chromosome strand, Stargazer resolves the haplotype by attempting to assign as many variants that define a single haplotype with any remaining variants being assigned to the second haplotype (i.e 4:1 is preferred over 3:2). PharmCAT instead provides all possible genotypes for the given set of variants. If PharmCAT identifies a combination of variants that do not translate to a known allele definition, the genotype is not called for the sample. Stargazer attempts to call any allele that is possible with the given set of variants, otherwise the sample is assigned the reference allele. Lastly, Stargazer uses Beagle as part of its pipeline which can impute missing genotype calls that are identified with high accuracy, although this occurrence has been found to be rare (<50 genotype calls). Overall, our data show that these differing approaches rarely resulted in differing calls and therefore unlikely had a significant impact on the frequencies reported.

The clinical impact of PGx is clear as where 1 in 5 of participants had an actionable phenotype and previous exposure to a medication with CPIC or FDA PGx prescribing guidance for that phenotype. Despite the high prevalence of PGx medication exposure events, the observed prevalence in *All of Us* is likely an underestimate since *All of Us* captures primarily inpatient medication exposures, all topical products were excluded, and important pharmacogenes (e.g. *CYP2D6, HLA-A, HLA-B*) were not included in the current genetic analysis. Importantly, the prevalence of PGx medication exposure events were found to be higher than previously reported data from large health systems(*11, 13, 14*). Similarly, prevalence of participants who have had an exposure to medications with associated actionable phenotypes were higher than previously reported rates in large biobank studies(*9, 14*). We attribute this difference for two reasons. First, CPIC continues to evaluate literature and update existing guidelines with addition of new medications(*23*) as well as developing new guidelines(*24, 25*). Some or all of these guidelines were not available when previous studies were conducted and therefore resulted in prevalence rates that were lower than what was observed within the current data from *All of Us*. Further, these studies were limited to an integrated call set consisting of WES and imputed array compared to WGS in *All of Us* and are unable to evaluate all known PGx variants.

### Limitations

The analysis focused on all genes that were part of a CPIC Level A gene-drug pair, but had notable exclusions (e.g. *CYP2D6*, *HLA-A, HLA-B*, and *MT-RNR1)*. *MT-RNR1* is within mitochondrial DNA, which was not available at the time of this study. *CYP2D6* and the HLAs were excluded because BAM/CRAM files were not yet available at the time of this study. Similarly, *CYP2C19* (*36, *37) and *SLCO1B1* (*48, *49) both have structural variants, however these are expected to be rare and could not be not characterized at this time. During haplotype calling, variants were excluded in *NUDT15* and *SLCO1B1* in addition to *CYP2B6* as a whole. This limited characterization of differences between a *12 and *1 in *NUDT15*, *45 and *1 in *SLCO1B1*, *46 and *15 in *SLCO1B1*. These alleles are expected to be rare based on overall variant frequencies within gnomAD (v3.1.2) at 0.00008 for NUDT15 *12 and 0.0016 for c.1738C>T defined for both *45 and *46 in *SLCO1B1*. *CYP2B6* is known to present a challenge in genotyping using short-read WGS due to the highly homologous pseudogene *CYP2B7* which has limited accuracy in characterizing variants(*26*). Further, while the *All of Us* dataset, provides an exceptional opportunity to link genetic, medication exposure, and phenotypes, medication exposure data are limited by whether a participant provided EHR data, and the amount of data contained in EHRs of the recruiting organization. Medication data may therefore be inconsistent between participants and may not include all medication exposures from each participant (e.g. may be missing over the counter medication exposures or some prescribing events when they occurred externally). Medication exposure events that included topical products or ambiguous formulations for a medication that could be topical were also excluded and represented roughly 6% of medication exposures. Overall, incomplete medication exposure information likely underestimated the true prevalence rate of medication exposure events that could be impacted by PGx. Lastly, while the *All of Us* cohort is diverse compared to other large datasets, it currently has low representation from some biogeographical groups in these early data releases. As *All of Us* achieves its goals to recruit participants that are traditionally underrepresented in research, we expect diversity will continue to expand and increase the value of these data.

## Data Availability

All data used in this analysis is available through the All of Us Researcher Workbench. The code is made available as a demonstration project within a featured workspace titled "Demo - Pharmacogenomics (PGx) variant frequency and medication exposures". Further, the PGx haplotype calls from both Stargazer and PharmCAT are available to all users for additional genotype-phenotype discovery and can be accessed through the featured workspace.

https://workbench.researchallofus.org/workspaces/aou-rw-87ca7327/demopharmacogenomicspgxvariantfrequencyandmedicationexposures/data

## Acknowledgments

The All of Us Research Program is supported (or funded) by the National Institutes of Health, Office of the Director: Regional Medical Centers: 1 OT2 OD026548; 1 OT2 OD026549; 1 OT2 OD026551;1 OT2 OD026552; 1 OT2 OD026553; 1 OT2 OD026554; 1 OT2 OD026555; 1 OT2 OD026556; IAA: AOD21037, AOD22003, AOD16037, AOD21041; Federally Qualified Health Centers: 75FCMC18D0047/75N98019F01202; Data and Research Center: 1 OT2 OD35404; Biobank: 4 U24 OD023121; The Participant Center: 5 U24 OD023176; Participant Technology Systems Center: 1 OT2 OD030043; Genome Centers: 1 OT2 OD002748; 1 OT2 OD002750; 1 OT2 OD002751; 1 OT2 OD027070; Genetic Counseling Resource: 1OT2OD028251; and Communications and Engagement: 1 OT2 OD028422; 1 OT2 OD035980; 1 OT2 OD025276; 1 OT2 OD036185; 1 OT2 OD025277; 1 OT2 OD025315; 1 OT2 OD036098; 1 OT2 OD027077; 1 OT2 OD028404; 1 OT2 OD028414; 1 OT2 OD028395; 1 OT2 OD031918; 1 OT2 OD031932; 1 OT2 OD031919; 1 OT2 OD031925; 1 OT2 OD031915; 1 OT2 OD031916; 1 OT2 OD036485. In addition, the All of Us Research Program would not be possible without the partnership of its participants.

We thank our colleagues, Kelsey Mayo, Ashley Able, Ashley Green, and Sokny Lim for providing their support and input throughout the demonstration project lifecycle. We thank Jun Qian for providing input on the project’s code review. We thank Lee Lichtenstein and Jennifer Zhang for providing the data artifacts used for the project. We thank the All of Us Data and Research Center’s Research Support team for their help during implementation. We also thank the All of Us Science Committee and All of Us Steering Committee for their efforts evaluating and finalizing the approved demonstration projects. The All of Us Research Program would not be possible without the partnership of contributions made by its participants. See the supplementary information for a roster of past and present All of Us principal investigators. To learn more about the All of Us Research Program’s research data repository, please visit https://www.researchallofus.org/.

An exemption to the Data and Statistics Dissemination Policy has been granted by the All of Us Research Program Resource Access Board to report counts <20.

## Author contributions

Conceptualization: AH, PE, EV

Methodology: AH, AR, SM, JS, JK, MW, KP, KW, DK, SK, QW, EV, PE

Writing – original draft: AH, PE

Writing – review & editing: AH, AR, SM, JS, JK, EV, MW, KP, KW, DK, SK, QW, RG, GJ, JS, AM, AHR, JD, PE

## References and Notes

1. J. C. Denny, J. L. Rutter, D. B. Goldstein, A. Philippakis, J. W. Smoller, G. Jenkins, E. Dishman, The “All of Us” Research Program. N Engl J Med 381, 668–676 (2019).

2. M. V. Relling, T. E. Klein, CPIC: Clinical Pharmacogenetics Implementation Consortium of the Pharmacogenomics Research Network. Clin Pharmacol Ther 89, 464–467 (2011).

3. L. H. Cavallari, C. R. Lee, A. L. Beitelshees, R. M. Cooper-DeHoff, J. D. Duarte, D. Voora, S. E. Kimmel, C. W. McDonough, Y. Gong, C. V. Dave, V. M. Pratt, T. D. Alestock, R. D. Anderson, J. Alsip, A. K. Ardati, B. C. Brott, L. Brown, S. Chumnumwat, M. J. Clare-Salzler, J. C. Coons, J. C. Denny, C. Dillon, A. R. Elsey, I. S. Hamadeh, S. Harada, W. B. Hillegass, L. Hines, R. B. Horenstein, L. A. Howell, L. J. B. Jeng, M. D. Kelemen, Y. M. Lee, O. Magvanjav, M. Montasser, D. R. Nelson, E. A. Nutescu, D. C. Nwaba, R. E. Pakyz, K. Palmer, J. F. Peterson, T. I. Pollin, A. H. Quinn, S. W. Robinson, J. Schub, T. C. Skaar, D. M. Smith, V. B. Sriramoju, P. Starostik, T. P. Stys, J. M. Stevenson, N. Varunok, M. R. Vesely, D. T. Wake, K. E. Weck, K. W. Weitzel, R. A. Wilke, J. Willig, R. Y. Zhao, R. P. Kreutz, G. A. Stouffer, P. E. Empey, N. A. Limdi, A. R. Shuldiner, A. G. Winterstein, J. A. Johnson, I. Network, Multisite Investigation of Outcomes With Implementation of CYP2C19 Genotype-Guided Antiplatelet Therapy After Percutaneous Coronary Intervention. JACC Cardiovasc Interv 11, 181–191 (2018).

4. D. M. F. Claassens, G. J. A. Vos, T. O. Bergmeijer, R. S. Hermanides, A. W. J. van ’t Hof, P. van der Harst, E. Barbato, C. Morisco, R. M. Tjon Joe Gin, F. W. Asselbergs, A. Mosterd, J. R. Herrman, W. J. M. Dewilde, P. W. A. Janssen, J. C. Kelder, M. J. Postma, A. de Boer, C. Boersma, V. H. M. Deneer, J. M. Ten Berg, A Genotype-Guided Strategy for Oral P2Y(12) Inhibitors in Primary PCI. N Engl J Med 381, 1621–1631 (2019).

5. J. J. Swen, C. H. van der Wouden, L. E. Manson, H. Abdullah-Koolmees, K. Blagec, T. Blagus, S. Bohringer, A. Cambon-Thomsen, E. Cecchin, K. C. Cheung, V. H. Deneer, M. Dupui, M. Ingelman-Sundberg, S. Jonsson, C. Joefield-Roka, K. S. Just, M. O. Karlsson, L. Konta, R. Koopmann, M. Kriek, T. Lehr, C. Mitropoulou, E. Rial-Sebbag, V. Rollinson, R. Roncato, M. Samwald, E. Schaeffeler, M. Skokou, M. Schwab, D. Steinberger, J. C. Stingl, R. Tremmel, R. M. Turner, M. H. van Rhenen, C. L. Davila Fajardo, V. Dolzan, G. P. Patrinos, M. Pirmohamed, G. Sunder-Plassmann, G. Toffoli, H. J. Guchelaar, C. Ubiquitous Pharmacogenomics, A 12-gene pharmacogenetic panel to prevent adverse drug reactions: an open-label, multicentre, controlled, cluster-randomised crossover implementation study. Lancet 401, 347–356 (2023).

6. M. C. Mills, C. Rahal, The GWAS Diversity Monitor tracks diversity by disease in real time. Nat Genet 52, 242–243 (2020).

7. S. Fatumo, T. Chikowore, A. Choudhury, M. Ayub, A. R. Martin, K. Kuchenbaecker, A roadmap to increase diversity in genomic studies. Nat Med 28, 243–250 (2022).

8. S. Reisberg, K. Krebs, M. Lepamets, M. Kals, R. Magi, K. Metsalu, V. M. Lauschke, J. Vilo, L. Milani, Translating genotype data of 44,000 biobank participants into clinical pharmacogenetic recommendations: challenges and solutions. Genet Med 21, 1345–1354 (2019).

9. G. McInnes, A. Lavertu, K. Sangkuhl, T. E. Klein, M. Whirl-Carrillo, R. B. Altman, Pharmacogenetics at Scale: An Analysis of the UK Biobank. Clin Pharmacol Ther 109, 1528–1537 (2021).

10. M. Samwald, H. Xu, K. Blagec, P. E. Empey, D. C. Malone, S. M. Ahmed, P. Ryan, S. Hofer, R. D. Boyce, Incidence of Exposure of Patients in the United States to Multiple Drugs for Which Pharmacogenomic Guidelines Are Available. PLoS One 11, e0164972 (2016).

11. C. Chanfreau-Coffinier, L. E. Hull, J. A. Lynch, S. L. DuVall, S. M. Damrauer, F. E. Cunningham, B. F. Voight, M. E. Matheny, D. W. Oslin, M. S. Icardi, S. Tuteja, Projected Prevalence of Actionable Pharmacogenetic Variants and Level A Drugs Prescribed Among US Veterans Health Administration Pharmacy Users. JAMA Netw Open 2, e195345 (2019).

12. L. B. Ramsey, H. H. Ong, J. S. Schildcrout, Y. Shi, L. A. Tang, J. K. Hicks, N. El Rouby, L. H. Cavallari, S. Tuteja, C. L. Aquilante, A. L. Beitelshees, D. L. Lemkin, K. V. Blake, H. Williams, J. J. Cimino, B. H. Davis, N. A. Limdi, P. E. Empey, C. M. Horvat, D. P. Kao, G. P. Lipori, M. B. Rosenman, T. C. Skaar, E. Teal, A. G. Winterstein, A. Owusu Obeng, D. Salyakina, A. Gupta, J. Gruber, J. McCafferty-Fernandez, J. R. Bishop, Z. Rivers, A. Benner, B. Tamraz, J. Long-Boyle, J. F. Peterson, S. L. Van Driest, I. P. W. Group, Prescribing Prevalence of Medications With Potential Genotype-Guided Dosing in Pediatric Patients. JAMA Netw Open 3, e2029411 (2020).

13. J. K. Hicks, N. El Rouby, H. H. Ong, J. S. Schildcrout, L. B. Ramsey, Y. Shi, L. Anne Tang, C. L. Aquilante, A. L. Beitelshees, K. V. Blake, J. J. Cimino, B. H. Davis, P. E. Empey, D. P. Kao, D. L. Lemkin, N. A. Limdi, P. L. G, M. B. Rosenman, T. C. Skaar, E. Teal, S. Tuteja, L. K. Wiley, H. Williams, A. G. Winterstein, S. L. Van Driest, L. H. Cavallari, J. F. Peterson, I. P. W. Group, Opportunity for Genotype-Guided Prescribing Among Adult Patients in 11 US Health Systems. Clin Pharmacol Ther 110, 179–188 (2021).

14. S. S. Verma, K. Keat, B. Li, G. Hoffecker, M. Risman, C. Regeneron Genetics, K. Sangkuhl, M. Whirl-Carrillo, S. Dudek, A. Verma, T. E. Klein, M. D. Ritchie, S. Tuteja, Evaluating the frequency and the impact of pharmacogenetic alleles in an ancestrally diverse Biobank population. J Transl Med 20, 550 (2022).

15. B. Li, K. Sangkuhl, R. Whaley, M. Woon, K. Keat, M. Whirl-Carrillo, M. D. Ritchie, T. E. Klein, Frequencies of pharmacogenomic alleles across biogeographic groups in a large-scale biobank. Am J Hum Genet, (2023).

16. S. M. Caspar, T. Schneider, J. Meienberg, G. Matyas, Added Value of Clinical Sequencing: WGS-Based Profiling of Pharmacogenes. Int J Mol Sci 21, (2020).

17. U. Amstutz, L. M. Henricks, S. M. Offer, J. Barbarino, J. H. M. Schellens, J. J. Swen, T. E. Klein, H. L. McLeod, K. E. Caudle, R. B. Diasio, M. Schwab, Clinical Pharmacogenetics Implementation Consortium (CPIC) Guideline for Dihydropyrimidine Dehydrogenase Genotype and Fluoropyrimidine Dosing: 2017 Update. Clin Pharmacol Ther 103, 210–216 (2018).

18. D. Twesigomwe, B. I. Drogemoller, G. E. B. Wright, A. Siddiqui, J. da Rocha, Z. Lombard, S. Hazelhurst, StellarPGx: A Nextflow Pipeline for Calling Star Alleles in Cytochrome P450 Genes. Clin Pharmacol Ther 110, 741–749 (2021).

19. R. S. Gammal, M. H. Court, C. E. Haidar, O. F. Iwuchukwu, A. H. Gaur, M. Alvarellos, C. Guillemette, J. L. Lennox, M. Whirl-Carrillo, S. S. Brummel, M. J. Ratain, T. E. Klein, B. R. Schackman, K. E. Caudle, D. W. Haas, C. Clinical Pharmacogenetics Implementation, Clinical Pharmacogenetics Implementation Consortium (CPIC) Guideline for UGT1A1 and Atazanavir Prescribing. Clin Pharmacol Ther 99, 363–369 (2016).

20. M. L. Appell, J. Berg, J. Duley, W. E. Evans, M. A. Kennedy, L. Lennard, T. Marinaki, H. L. McLeod, M. V. Relling, E. Schaeffeler, M. Schwab, R. Weinshilboum, A. E. Yeoh, E. M. McDonagh, J. M. Hebert, T. E. Klein, S. A. Coulthard, Nomenclature for alleles of the thiopurine methyltransferase gene. Pharmacogenet Genomics 23, 242–248 (2013).

21. V. M. Pratt, W. Y. Wang, E. C. Boone, U. Broeckel, N. Cody, L. Edelmann, A. Gaedigk, T. C. Lynnes, E. B. Medeiros, A. M. Moyer, M. W. Mitchell, S. A. Scott, P. Starostik, A. Turner, L. V. Kalman, Characterization of Reference Materials for TPMT and NUDT15: A GeT-RM Collaborative Project. J Mol Diagn 24, 1079–1088 (2022).

22. A. Gaedigk, M. Ingelman-Sundberg, N. A. Miller, J. S. Leeder, M. Whirl-Carrillo, T. E. Klein, C. PharmVar Steering, The Pharmacogene Variation (PharmVar) Consortium: Incorporation of the Human Cytochrome P450 (CYP) Allele Nomenclature Database. Clin Pharmacol Ther 103, 399–401 (2018).

23. R. M. Cooper-DeHoff, M. Niemi, L. B. Ramsey, J. A. Luzum, E. K. Tarkiainen, R. J. Straka, L. Gong, S. Tuteja, R. A. Wilke, M. Wadelius, E. A. Larson, D. M. Roden, T. E. Klein, S. W. Yee, R. M. Krauss, R. M. Turner, L. Palaniappan, A. Gaedigk, K. M. Giacomini, K. E. Caudle, D. Voora, The Clinical Pharmacogenetics Implementation Consortium Guideline for SLCO1B1, ABCG2, and CYP2C9 genotypes and Statin-Associated Musculoskeletal Symptoms. Clin Pharmacol Ther 111, 1007–1021 (2022).

24. K. N. Theken, C. R. Lee, L. Gong, K. E. Caudle, C. M. Formea, A. Gaedigk, T. E. Klein, J. A. G. Agundez, T. Grosser, Clinical Pharmacogenetics Implementation Consortium Guideline (CPIC) for CYP2C9 and Nonsteroidal Anti-Inflammatory Drugs. Clin Pharmacol Ther 108, 191–200 (2020).

25. J. J. Lima, C. D. Thomas, J. Barbarino, Z. Desta, S. L. Van Driest, N. El Rouby, J. A. Johnson, L. H. Cavallari, V. Shakhnovich, D. L. Thacker, S. A. Scott, M. Schwab, C. R. S. Uppugunduri, C. M. Formea, J. P. Franciosi, K. Sangkuhl, A. Gaedigk, T. E. Klein, R. S. Gammal, T. Furuta, Clinical Pharmacogenetics Implementation Consortium (CPIC) Guideline for CYP2C19 and Proton Pump Inhibitor Dosing. Clin Pharmacol Ther 109, 1417–1423 (2021).

26. Z. Desta, A. El-Boraie, L. Gong, A. A. Somogyi, V. M. Lauschke, C. Dandara, K. Klein, N. A. Miller, T. E. Klein, R. F. Tyndale, M. Whirl-Carrillo, A. Gaedigk, PharmVar GeneFocus: CYP2B6. Clin Pharmacol Ther 110, 82–97 (2021).

27. A. H. Ramirez, L. Sulieman, D. J. Schlueter, A. Halvorson, J. Qian, F. Ratsimbazafy, R. Loperena, K. Mayo, M. Basford, N. Deflaux, K. N. Muthuraman, K. Natarajan, A. Kho, H. Xu, C. Wilkins, H. Anton-Culver, E. Boerwinkle, M. Cicek, C. R. Clark, E. Cohn, L. Ohno-Machado, S. D. Schully, B. K. Ahmedani, M. Argos, R. M. Cronin, C. O’Donnell, M. Fouad, D. B. Goldstein, P. Greenland, S. J. Hebbring, E. W. Karlson, P. Khatri, B. Korf, J. W. Smoller, S. Sodeke, J. Wilbanks, J. Hentges, S. Mockrin, C. Lunt, S. A. Devaney, K. Gebo, J. C. Denny, R. J. Carroll, D. Glazer, P. A. Harris, G. Hripcsak, A. Philippakis, D. M. Roden, P. All of Us Research, The All of Us Research Program: Data quality, utility, and diversity. Patterns (N Y) 3, 100570 (2022).

28. I. All of Us Research Program Genomics, Genomic data in the All of Us Research Program. Nature, (2024).

29. E. Venner, D. Muzny, J. D. Smith, K. Walker, C. L. Neben, C. M. Lockwood, P. E. Empey, G. A. Metcalf, C. Kachulis, G. All of Us Research Program Regulatory Working, S. Mian, A. Musick, H. L. Rehm, S. Harrison, S. Gabriel, R. A. Gibbs, D. Nickerson, A. Y. Zhou, K. Doheny, B. Ozenberger, S. E. Topper, N. J. Lennon, Whole-genome sequencing as an investigational device for return of hereditary disease risk and pharmacogenomic results as part of the All of Us Research Program. Genome Med 14, 34 (2022).

30. K. J. Karczewski, L. C. Francioli, G. Tiao, B. B. Cummings, J. Alfoldi, Q. Wang, R. L. Collins, K. M. Laricchia, A. Ganna, D. P. Birnbaum, L. D. Gauthier, H. Brand, M. Solomonson, N. A. Watts, D. Rhodes, M. Singer-Berk, E. M. England, E. G. Seaby, J. A. Kosmicki, R. K. Walters, K. Tashman, Y. Farjoun, E. Banks, T. Poterba, A. Wang, C. Seed, N. Whiffin, J. X. Chong, K. E. Samocha, E. Pierce-Hoffman, Z. Zappala, A. H. O’Donnell-Luria, E. V. Minikel, B. Weisburd, M. Lek, J. S. Ware, C. Vittal, I. M. Armean, L. Bergelson, K. Cibulskis, K. M. Connolly, M. Covarrubias, S. Donnelly, S. Ferriera, S. Gabriel, J. Gentry, N. Gupta, T. Jeandet, D. Kaplan, C. Llanwarne, R. Munshi, S. Novod, N. Petrillo, D. Roazen, V. Ruano-Rubio, A. Saltzman, M. Schleicher, J. Soto, K. Tibbetts, C. Tolonen, G. Wade, M. E. Talkowski, C. Genome Aggregation Database, B. M. Neale, M. J. Daly, D. G. MacArthur, The mutational constraint spectrum quantified from variation in 141,456 humans. Nature 581, 434–443 (2020).

31. V. M. Pratt, L. H. Cavallari, A. L. Del Tredici, H. Hachad, Y. Ji, L. V. Kalman, R. C. Ly, A. M. Moyer, S. A. Scott, M. Whirl-Carrillo, K. E. Weck, Recommendations for Clinical Warfarin Genotyping Allele Selection: A Report of the Association for Molecular Pathology and the College of American Pathologists. J Mol Diagn 22, 847–859 (2020).

32. A. o. U. Genome Centers, A. o. U. Data and Research Center, “Genomic Research Data Quality Report,” (https://support.researchallofus.org/hc/en-us/articles/14969499850644-All-of-Us-Genomic-Quality-Report-ARCHIVED-C2022Q2R2-CDR-CT-Dataset-v6-, 2022).

33. S. B. Lee, M. M. Wheeler, K. E. Thummel, D. A. Nickerson, Calling Star Alleles With Stargazer in 28 Pharmacogenes With Whole Genome Sequences. Clin Pharmacol Ther 106, 1328–1337 (2019).

34. K. Sangkuhl, M. Whirl-Carrillo, R. M. Whaley, M. Woon, A. Lavertu, R. B. Altman, L. Carter, A. Verma, M. D. Ritchie, T. E. Klein, Pharmacogenomics Clinical Annotation Tool (PharmCAT). Clin Pharmacol Ther 107, 203–210 (2020).

35. K. E. Caudle, H. M. Dunnenberger, R. R. Freimuth, J. F. Peterson, J. D. Burlison, M. Whirl-Carrillo, S. A. Scott, H. L. Rehm, M. S. Williams, T. E. Klein, M. V. Relling, J. M. Hoffman, Standardizing terms for clinical pharmacogenetic test results: consensus terms from the Clinical Pharmacogenetics Implementation Consortium (CPIC). Genet Med 19, 215–223 (2017).

36. R. Huddart, A. E. Fohner, M. Whirl-Carrillo, G. L. Wojcik, C. R. Gignoux, A. B. Popejoy, C. D. Bustamante, R. B. Altman, T. E. Klein, Standardized Biogeographic Grouping System for Annotating Populations in Pharmacogenetic Research. Clin Pharmacol Ther 105, 1256–1262 (2019).

37. FDA. (https://www.fda.gov/medical-devices/precision-medicine/table-pharmacogenetic-associations, 2022).

