## Supplement for "Frequency of pharmacogenomic variation and medication exposures among All of Us Participants"

**The PDF file includes:**

Materials and Methods

Tables S1 to S9 captions

All of Us Research Program Investigators author list

Materials and Methods

***Experimental Design***

The objective of this study was to compute all known allele and phenotype frequencies for PGx genes among diverse *All of Us* participants. Short read WGS data (n=98,590) was used to extract all known variants for 16 PGx genes. Genetic data quality control metrics included variant call rate, sample call rate, and deviation from Hardy-Weinburg equilibrium (HWE). PGx haplotype calling was completed using Stargazer and PharmCAT as an orthogonal method. Predicted phenotypes were manually assigned using CPIC defined terms. Computed variant, allele, and phenotype frequencies were compared to gold standard databases in gnomAD and CPIC. Finally, medication exposures were extracted from EHR data using RxNorm IDs to evaluate the projected impact of PGx by computing the frequency of participants who had a predicted phenotype that has a recommendation for a change in therapy along with an affected medication exposure.

***Data access through the All of Us Researcher Workbench***

This research was an approved *All of Us* demonstration project proposed by Consortium members, reviewed and overseen by the program, and confirmed as meeting criteria for non-human subjects research by the *All of Us* Institutional Review Board.

*All of Us* recruitment methods, sites, and data have been described previously(*1, 27*). Participant surveys, electronic health records (EHR), and genomic data are accessible using the *All of Us* Researcher Workbench (https://www.researchallofus.org/), a cloud-based platform where approved researchers can access and analyze *All of Us* data. The Researcher Workbench currently offers tools with a user interface built for creating datasets for analysis (Dataset Builder) and Workspaces within Jupyter Notebooks to analyze data using R and Python programming languages. EHR data was provided and linked for participants who consented to make their data available. EHR data types are mapped to the Observational Medicines Outcomes Partnership (OMOP) common data model v5.2 maintained by the Observational Health Data Sciences and Informatics collaborative.

***Genomic data***

Genomic data analyzed included short-read WGS (n=98,590) and genotyping array (n=165,127) from the v6 Curated Data Repository of the *All of Us* cohort(*28*). Briefly, short-read WGS and array genotyping was completed at three genomic centers (Baylor College of Medicine, The Broad Institute, and the Northwest Genomics Center at the University of Washington). For WGS, DRAGEN v3.4.12 (Illumina, San Diego, CA) was used to compute quality metrics and alignment to the GRCh38 reference genome with variant level data made available as a hail MatrixTable(*29*). Array genotyping was completed using the Infinium™ Global Diversity Array (Illumina, San Diego, CA). All genetic data excluded participants who have previously received an allogeneic bone marrow transplant as identified through the EHR data but included participants deemed to have any degree of genetic relatedness.

***Genetic ancestry***

Genetic ancestry was computed using principal component analysis by the *All of Us*-DRC(*28*). High quality exon sites (n=151,159) from all autosomes were selected in both the training dataset (Human Genome Diversity Project + 1000 Genomes) and the test dataset (WGS) to generate the first sixteen principle components (PCs). The PCs were then used as features for a random forest classifier. Samples were assigned ancestry grouping if the probability was >90%, otherwise they were assigned “Other”. Ancestry super-populations used gnomAD definitions which include African/African American (AFR), American Admixed/Latino (AMR), East Asian (EAS), European (EUR), Middle Eastern (MID), South Asian (SAS), and Other (OTH) (*30*).

***Extraction of PGx variants from WGS data***

Sixteen pharmacogenes and gene regions that are part of a CPIC level A gene-drug pair with guideline evidence for a drug association were included in the analysis (*ABCG2, CACNA1S, CYP2B6, CYP2C Cluster, CYP2C19, CYP2C9, CYP3A5, CYP4F2, DPYD, G6PD, NUDT15, RYR1, SLCO1B1, TPMT, UGT1A1, VKORC1)*. *CYP2D6*, *HLA-A,* and *HLA-B* were not analyzed at this time as read alignment files were not yet available. *MT-RNR1* was also excluded due to lack of mitochondrial DNA data at this time. Targeted variants (n=955) included all variants present in PharmVar (version 5.2.6), CPIC (accessed June 21, 2022), and the Tier 1 and 2 tables within the Association for Molecular Pathology (AMP) guidelines for the selected genes and gene regions (table S1) (*31*). Genotype calls were extracted from the WGS hail MatrixTable for PGx variants as a VCF file. Positions missing from the complete dataset were assumed to be homozygous reference for all samples. Any variants labeled as low quality based on the *All of Us* Genomic Research Data Quality Report were excluded(*32*). Variant and sample call rate were computed with cutoffs of 0.98 and 0.95 respectively. Exact test of Hardy-Weinberg equilibrium (HWE) was computed for all biallelic non-chromosome X variants with a p-value cutoff of 2.15x10^-5^ (n=2,331 tests). Sex was imputed as part of the DRAGEN pipeline and used for *G6PD* genotype assignment.

***PGx haplotype calling***

PGx haplotype calling was completed using both Stargazer v2.0(*33*) and PharmCAT v2.2.1(*34*) as an orthogonal confirmation. Both tools have been previously validated against a set of GeT-RM samples (*15, 33*). The following assumptions are made in both calling pipelines: 1) input genetic data was unphased; 2) if a distinction between >1 genotype calls cannot be made or if a sample was found to have >2 alleles, an “Indeterminate/Indeterminate” genotype is assigned (for *G6PD* XY samples, “Indeterminate” genotype when there was >1 allele). *DPYD* allele translations were based on the alleles defined by CPIC. *DPYD* genotypes were assigned regardless of the number of alleles present with activity score calculated based on the two identified alleles with the lowest function as described in the CPIC guideline(*17*). *G6PD* XY samples were assigned a single allele as the genotype, while samples that were XX or unknown were assigned a diplotype. Predicted phenotypes were manually assigned using the CPIC API Predicted phenotypes for *CYP2C Cluster*, *VKORC1*, and *CYP4F2* were assigned “Variant Present'' or “Variant Absent” based on the presence of the rs12777823, c.-1639G>A, and c.1297G>A (*3) variants respectively as standardized phenotypes are not currently defined by CPIC(*35*).

***Variant, allele, and predicted phenotype frequencies***

Variant and allele frequencies were calculated as the number of variant calls/alleles divided by the total number of available calls/alleles (excludes sample with no calls for variants). For *G6PD,* frequency calculations excluded samples not assigned XX or XY due to lack of information on chromosome X ploidy. For *DPYD*, allele frequencies were computed independently as samples can carry >2 alleles due to presence of haplotype blocks and not true haplotypes(*17*).

Variant frequencies across genetic ancestries were compared to gnomAD v3.1.2 (API accessed Jan. 14, 2023)(*30*). Allele and phenotype frequencies were compared to CPIC across genetic ancestries mapped to PharmGKB biogeographical groups where appropriate (CPIC table S2, API accessed Nov 10, 2022)(*36*). An actionable phenotype was defined as a predicted phenotype for a gene or gene region which has a recommendation from CPIC or the FDA for a change in prescribing through either an alternative treatment option or a change in the dose(*37*). The “Variant Present” phenotype for *CYP4F2* and *CYP2C Cluster* was considered actionable independent of genetic ancestry.

***Demographics and medication exposures***

Participant demographics and medication exposures were extracted from *All of Us* controlled tier v6 cohort consisting of EHR data with a date cutoff of January 1st, 2022. Survey demographic data was used for reporting counts of self-reported sex at birth and gender in Table 1. All other analysis used genetic ancestry groups. A medication or drug exposure is defined using the OMOP v5.2 common data model which captures any event involving a medication. There were 70 medications identified with CPIC Level A evidence with a guideline (table S3; CPIC accessed March 28, 2023) with an additional 46 medications listed on the FDA Table of PGx Associations in sections 1 and 2 (Accessed March 28, 2023)(*37*). PGx medication exposures were identified based on the presence of a standard or source concept code involving medications. Exposures were filtered for the selected medications using RxNorm IDs followed by removal of false positives using medication generic and brand name. Secondary string-based filtering was performed to further identify medication exposure events using generic or brand name. Topical formulations or any formulation that was ambiguous for any medication that could have a topical product were excluded.

***Statistical Analysis***

Analysis was completed using Python (v3.7.12) within Jupyter Notebooks using hail (v0.2.107), pandas (v1.3.5), scipy (v1.7.3), and numpy (v1.21.6) libraries. Figures were created using the matplotlib (v3.5.1) library. All code has been made available for re-use on the *All of Us* researcher workbench as a featured workspace along with the PGx haplotype calls completed using Stargazer and PharmCAT. Variant, allele, and phenotype frequencies by genetic ancestry comparisons to gnomAD and CPIC were completed using Fisher’s exact test or Chi squared where appropriate with p-value cutoffs of 5.04x10^-5^ (n=992 tests), 1.16x10^-4^ (n=431 tests), and 2.45x10^-4^ (n=204 tests) respectively.

Supplementary Table 1: Variant Frequencies. Variants presented represent all variants for the fifteen CPIC-A genes obtained from PharmVar, CPIC, and Tier 1 and Tier 2 AMP Guideline tables. *CYP2B6* and any variants dropped due to low quality are excluded. Hardy-Weinburg equilibrium was computed for all biallelic non-chromosome X variants. Values that were not calculated represent variants that were not present in the dataset where all samples are homozygous reference.

Supplementary Table 2: Genetic Ancestry CPIC Mapping. The mapping represents approximate overlap between imputed genetic ancestry superpopulations and CPIC biogeographical groups.

Supplementary Table 3: Medications. Medications identified with CPIC level A evidence with an associated gene(s) or from FDA Tables of Pharmacogenetic Associations 1 and 2 (CPIC accessed June 21, 2022).

Supplementary Table 4: Allele Frequencies. Allele frequencies overall and by genetic ancestry computed for Stargazer and PharmCAT. Structural variants (CYP2C19 *36 and *37; SLCO1B1 *48 and *49) were not characterized. Alleles frequencies related to dropped variants (NUDT15 *12 and *1; SLCO1B1 *45 and *1; SLCO1B1 *46 and *15) cannot be distinguished at this time.

Supplementary Table 5: Phenotype Frequencies. Phenotype frequencies overall and by genetic ancestry computed for Stargazer and PharmCAT. Predicted phenotypes for *CYP2C Cluster*, *VKORC1*, and *CYP4F2* were assigned “Variant Present” or “Variant Absent” based on the presence of the rs12777823, c.-1639G>A, and c.1297G>A (*3) variants respectively.

Supplementary Table 6: Genotypes with many alleles. Genotypes were identified where a sample carried >2 alleles for a gene (>1 for *G6PD* XY samples). Multiple possible genotypes are semicolon delimited when there is phasing ambiguity.

Supplementary Table 7: Frequency comparison to known databases. Computed variant, allele, and phenotype frequencies compared to known frequencies in gnomAD and CPIC. Values represent the number of variant/allele/phenotypes by genetic ancestry which are similar or significantly different than known values.

^*^Compared to gnomAD v3.1.2 across genetic ancestry groups (excludes OTH)

^†^Compared to the overlapping biogeographical groups within CPIC as shown in Supplementary Table 2.

^‡^Reference alleles, the other genetic ancestry group, *1 (former reference allele) in *CYP2C19*, *RYR1*, and *CACNA1S* were excluded from comparison.

Supplementary Table 8: New allele and phenotype frequencies. Counts of allele and phenotype frequencies by genetic ancestry that were previously unknown or reported as 0 that were quantified during this study.

Supplementary Table 9: Overall medication prevalence. The overall prevalence determined from EHR medication exposure data for all medications with CPIC level A evidence or are present within FDA Table of PGx Associations in sections 1 and 2 (CPIC accessed June 21, 2022).

Acknowledgement List of Principal Investigators

Past and Present All of Us Research Program Principal Investigators

^1^Dmitry Abramov; ^2^Brian Ahmedani; ^3^Habib Ahsan; ^4^Hoda Anton-Culver; ^5^Maria Argos; ^6^Dave W. Arnold; ^5^Brisa Aschebook-Kilfoy; ^7^Katie Baca-Motes; ^8^Catherine Balthazar; ^9^Laura Bartlett; ^6^Gillian Bartlett-Esquilant; ^10^Melissa Basford; ^11^Leland Baskin; ^12^Mark Begale; ^13^Eric Boerwinkle; ^14^Ingrid Bonilla-Mercado; ^15^Alexander Borowsky; ^16^Amaryllis Silva Boschetti; ^17^Matthew Breeden; ^18^Brad Bryan; ^19^Jessica Burke; ^20^Elizabeth Burnside; ^21^Lisa Cadmus-Bertram; ^22^Librada Callender; ^23^Roberta Carlin; ^24^Olveen Carrasquillo; ^25^Suchitra Chandrasekaran; ^26^Alexander Charney; ^27^Mine Cicek; ^28^Cheryl Clark; ^29^Elizabeth Cohn; ^30^Vivian Colon-Lopez; ^31^Alejandro Comellas; ^23^Karl Cooper; ^32^Linda Cottler; ^33^Errol Crook; ^34^Elizabeth Culler; ^5^Martha Daviglus; ^35^Kathy Deerinwater; ^36^Liliana Lombardi Desa; ^37^Jacob Ditsch; ^38^Kim Doheny; ^23^Charles Drum; ^32^Milton Eder; ^39^Mark Edmunds; ^40^Evan Eichler; ^41^Leslie Eiland; ^17^Kim Enard; ^42^Rachel Everhart; ^43^Adolph Falcon; ^20^Dorothy Farrar-Edwards; ^44^Becky Fein; ^45^Zeno Frano; ^46^Yuri Fresko; ^47^Gretchen Funk; ^48^Stacey Gabriel; ^49^Marie Gantz; ^50^Michael Garrett; ^51^Ali G Gharavi; ^52^Richard Gibbs; ^53^David Glazer; ^54^Marc Goodman; ^55^Philip Greenland; ^56^Allen Greiner; ^17^Richard Grucza; ^14^Lourdes Guerrios-Rivera; ^56^Aditi Gupta; ^57^Sandra Halverson; ^58^Eileen Handberg; ^10+^Paul Harris; ^59^Scott Joseph Hebbring; ^60^Robert Hiatt; ^55^Joyce Ho; ^58^William R Hogan; ^61^Beverly Wilson Holmes; ^44^Laura Horne; ^51^George Hripcsak; ^62^Priscilla Igho-Pemu; ^24^Rosario Isasi; ^63^Jessica Isom; ^12^Neeta Jain; ^12^Praduman Jain; ^64^Jessica Jarmin; ^40^Gail Jarvik; ^2^Christine D Cole Johnson; ^65^Megan Jula; ^66^Royan Kamyar; ^67^Elizabeth W Karlson; ^68^Rainu Kaushal; ^41^Amber Brown Keebler; ^25^Robert Kelley; ^36^Kathleen Keogh; ^69^Vik Kheterpal; ^70^Sue Kim; ^29^Frida Kleiman; ^71^Isaac Kohane; ^72^Bruce Korf; ^73^Monica Kraft; ^50^Babbette Lamarca; ^74^Linda Perez Laras; ^52^Brendan Lee; ^48^Niall Lennon; ^75^Dessie Levy; ^72^Beth Lewis; ^76^Elizabeth Cora Lewis; ^77^Megan Lewis; ^78^David Ming-Hung Lin; ^79^Christopher Lough; ^80^Mitchell Lunn; ^12^David Klein, MBA; ^81^Todd Mahr; ^82^Emily Makahi; ^17^Ted Malmstrom; ^83^Oscar Marroquin; ^84^Judith Martin; ^85^Vivienne Marshall;Elizabeth Mayer-Davis; ^24^Jacob McCauley; ^86^Brinkoetter April McCrea; ^86^Patrick McGovern; ^87^Jeffrey McKinney; ^13^David McPherson; ^88^Daniella Meeker; ^62^Robert Meller; ^30^Jose Melo; ^75^Michael Minor; ^14^Irma Molina; ^88^Janitza Montalvo-Ortiz; ^7^Julia Moore-Vogel; ^89^Francisco A Moreno; ^67^Shawn N Murphy; ^90^Dianne Debra Murray; ^7^Evan Muse; ^63^Paul Nemeskal; ^91^George T O'Connor; ^80^Juno Obedin-Maliver; ^16^Lucila Ohno-Machado; ^56^Akinlolu Ojo; ^56^Tammy Ojo; ^82^May Okihiro; ^25^Matt Pagel; ^92^Kapil Parakh; ^6^Elizabeth Parks; ^2^Cathryn Peltz-Rauchman; ^56^Jill Peltzer; ^18^Tim Peterson; ^48^Anthony Philippakis; ^93^Fornessa Randal; ^94^Subhara Raveendran; ^48^Heidi Rehm; ^95^Gail Reilly; ^46^Jody Reilly; ^96^Eric Reiman; ^83^Steven Reis; ^31^Heather Reisinger; ^84^Anne-Marie Rick; ^43^Edgar M Gil Rico; ^74^Nelida Rivera; ^8^Tonya Roberson; ^10^Dan Roden; ^52^Laura Rosales; ^68^Margaret Elizabeth Ross; ^25^Tracie Rosser; ^97^Mercedita Roxas-Murray; ^98^Bruce Sachais; ^56^Matthias Salathe; ^99^Linda Salgin; ^100^Sherilyn Sawyer; ^17^Jeffrey Scherrer; ^61^Eric Schlueter; ^101^A. Mark Schmidt; ^102^Nicole Seagriff; ^58^Elizabeth Shenkman; ^103^William Simonson; ^16^Amy Sitapati; ^59^Sanjay Skukla; ^104^Debra Smith; ^67^Jordan Smoller; ^63^Cynthia So-Armah; ^105^Bento Marcelo Soares; ^106^Arambula Teshia G. Solomon; ^31^Kimberly Sprenger; ^107^Louisa Stark; ^108^Gene Stegeman; ^12^Scott Sutherland; ^109^Christin Suver; ^110^Michael Taitel; ^99^Gregory Talavera; ^111^Amy Taylor; ^95^Kyla Taylor; ^112^Ronnie Tepp; ^96^Andreas A Theodorou; ^27^Steve Thibodeau; ^113^Cheryl Thomas; ^56^Jeffrey Thompson; ^95^Daniel Hernandez Tinoco; ^7^Eric Topol; ^114^Scott Topper; ^115^Rhonda Trousdale; ^116^Phil Tsao; ^117^Bryce Turner; ^77^Jen Uhrig; ^106^Celina Valencia; ^100^Jason Vassy; ^83^Shyam Visweswaran; ^25^Miriam Vos; ^6^Russ Waitman; ^87^Jamie Walz; ^118^Preston Watkins; ^119^Chia-Lin Wei; ^120^Jones Deborah Weiss; ^28^Scott T Weiss; ^45^Jeffrey Whittle; ^121^Blaker Wilkerson; ^10^Consuelo Wilkins; ^122^Steve Wimmer; ^95^Eboni Winford; ^111^Katrina Yamazaki; ^123^Fady Youssef; ^114^Alicia Zhou; ^24^Stephan Zuchner;

Note

This is the list of individuals who were Principal Investigators or equivalent with the *All of Us* Research Program during the period that this paper was in development, May 1, 2021 - June 1, 2024.

Legend

+ Principal Investigator/Lead Author for the *All of Us* Research Program protocol

Affiliations

^1^RMC AoUSCC: Loma Linda University Health

^2^Henry Ford Health System

^3^University of Chicago Medical Center

^4^University of California, Irvine

^5^University of Illinois at Chicago

^6^RMC Heartland: University of Missouri Medical Center

^7^Scripps Research Translational Institute

^8^RMC IPMC: Governor's State University

^9^National Library of Medicine (NLM)

^10^Vanderbilt University Medical Center

^11^TPC: Our Blood Institute

^12^Vibrent Health

^13^University of Texas Health Science Center at Houston

^14^VA: VA Caribbean Health Care

^15^University of California, Davis

^16^University of California, San Diego

^17^Saint Louis University

^18^TPC: BioLinked

^19^MITRE Corporation

^20^University of Wisconsin at Madison

^21^RMC Wisconsin: U Wisconsin Madison

^22^RMC TACH: Baylor Scott & White Health

^23^American Association of Health and Disability

^24^University of Miami School of Medicine

^25^Emory University

^26^RMC Arizona: Mount Siani Health System

^27^Mayo Clinic and Foundation, Rochester

^28^Brigham and Women's Hospital

^29^Hunter College

^30^University of Puerto Rico Comprehensive Cancer Center

^31^RMC Heartland: University of Iowa Hospitals & Clinics

^32^CTSA Community Engagement Programs

^33^University of South Alabama

^34^TPC: Blood Assurance

^35^AI/IN: American Indian Sceince and Engineering Society (AISES)

^36^Sun River Health

^37^CEP: Am Assn Health and Disability

^38^Johns Hopkins University School of Medicine

^39^San Diego Blood Bank

^40^University of Washington

^41^RMC Heartland: University of Nebraska Medical Center

^42^TPC: Denver Health

^43^National Alliance for Hispanic Health

^44^TPC: Active Minds

^45^Medical College of Wisconsin

^46^Quest Diagnostics Incorporated

^47^FiftyForward

^48^Broad Institute

^49^NPH: Research Triangle

^50^University of Mississippi Medical Center

^51^Columbia University

^52^Baylor University

^53^Verily Life Sciences

^54^Cedars Sinai

^55^Northwestern University

^56^RMC Heartland: University of Kansas Medical Center

^57^TPC: DLH Corp

^58^University of Florida

^59^Marshfield Clinic Research Institute

^60^University of California, San Francisco

^61^Cooperative Health

^62^Morehouse School of Medicine, Atlanta

^63^Mass General Hospital

^64^Tactis

^65^TPC: Mary’s Center

^66^TPC: Owaves

^67^Partners Health Care

^68^Cornell University, Weill Medical College

^69^CareEvolution, Inc.

^70^University of Southern California

^71^Harvard Medical School

^72^University of Alabama at Birmingham

^73^Mount Sinai Health System

^74^COSSMA

^75^National Baptist Convention

^76^RMC Southern: U Alabama, Birmingham (UAB)

^77^Research Triangle Institute

^78^TPC: Bloodworks Northwest

^79^LifeSouth

^80^Stanford University

^81^Gundersen Health System

^82^Waianae Coast CHC

^83^University of Pittsburgh

^84^RMC Pitt: U Pittsburgh

^85^South Texas Blood and Tissue Center

^86^Wondros

^87^Sensis

^88^Yale

^89^University of Arizona, Tucson

^90^CEP: Baylor College of Medicine

^91^Boston Medical Center

^92^TPC: Fitbit

^93^Asian Health Coalition

^94^Patients Like Me

^95^Cherokee Health Systems

^96^Banner Health

^97^Montage Marketing Group

^98^TPC: New York Blood Center Enterprises (NYBC)

^99^San Ysidro Health Center

^100^VA AoU Coordinating Center

^101^RMC TACH: Kaiser Permanente

^102^FQHC: Community Health Ctr

^103^Cascade Regional BLood Services

^104^SunCoast Blood Center

^105^RMC IPMC: UI Chicago Hosp & Health Sci Sys

^106^Developing Partnerships to Advance Precision Medicine with AI/AN Tribal Nations

^107^University of Utah

^108^ExamOne

^109^Sage Bionetworks

^110^Walgreen Co.

^111^Community Health Center, Inc.

^112^HCM Strategists

^113^Delta Research and Educational Foundation

^114^Color Genomics, Inc.

^115^NYC Health + Hospitals

^116^VA AoU Coordinating Center - Palo Alto

^117^University Southern California

^118^WebMD Health Corp

^119^Gen NW: U Washington

^120^RMC SouthEast: U Miami Sch of Med

^121^Blue Cross Blue Shield

^122^TPC: 1nHealth

^123^Memorial Health Services
